# Measuring the impact of lived experience and caregiver engagement in research on the research conducted: development and pilot testing of an assessment tool

**DOI:** 10.64898/2026.04.01.26349956

**Authors:** Lisa D. Hawke, Jingyi Hou, Katie Upham, Mary Rose van Kesteren, Charlotte Munro, Shoshana Hauer, Claudia Sendanyoye, Tanya Halsall, Lena C. Quilty, Clayon Hamilton, Skye Barbic, Wei Wang

## Abstract

**Background.:** People with lived/living experience of health conditions, as well as caregivers, are increasingly engaged in research. This study aimed to develop and pilot test a new tool measuring the impact of lived/living experience engagement on research. The measurement tool is called the *Measure of Engagement Tool for Research and lived Experience* (METRE).

**Method.:** We conducted a qualitative descriptive study among 28 people with lived/living experience and caregivers and 12 academic researchers to understand the impacts of engagement on the research. Using the findings, we drafted the METRE. We pilot tested the METRE among 13 people with lived/living experience and caregivers and 10 academic researchers. Insights were used to refine the METRE.

**Results.:** Qualitatively, participants identified multiple domains of impact of engagement on research, which guided tool development. Pilot testing of the draft METRE revealed it being straightforward to complete, while providing a thorough evaluation of the impact that engagement has on research. However, some areas of improvement were recommended. The draft items showed acceptable preliminary performance.

**Conclusions.:** An assessment tool is now available to assess the impact that lived/living experience engagement has on research. Additional research is required to evaluate its psychometric properties. Tools to evaluate the impact of engagement on research will help advance the science of engagement and support engaged research teams in their work.

## 1 Background

People with lived and living experience of health challenges, as well as family members or caregivers, are increasingly engaged in research [1, 2]. Lived/living experience partners are experts of their own experience in research about their needs [3]. This type of engagement is often referred to as ‘patient-engaged research,’ or ‘patient-oriented research [4]’, although lived/living experience is preferred by some populations [5]. Patient-oriented research practices call for people with lived/living experience and caregivers to be engaged in the research process.

Those with lived/living experience and caregivers can be engaged in research across study designs and research topics [1, 6, 7]. They can provide guidance on, collaborate on, co-design, and/or lead many aspects of research. These include setting research priorities, selecting methodologies, conducting research, analyzing and interpreting data, and co-creating and disseminating research knowledge [7]. Patient-engaged research approaches are strongly guided by pragmatism as a fundamental paradigm underlying the work, where research evidence and experiential knowledge are equally valued [8]. Engagement is an ethical imperative and anti-oppressive practice in response to past and ongoing inequities in research and clinical practice [9, 10]. The foundation of authentic engagement practices involves building a team that values the knowledge of lived/living experience and caregiver partners, in order to work collaboratively [8].

Despite the growing trend toward lived/living experience engagement in health research, measurement science has been identified as a gap in the science of engagement [11]. To our knowledge, there is currently no validated assessment tool available to measure the perceived impact of engagement on the research, i.e., how much the research project is changed based on their input. The Patient Engagement in Research Scale (PEIRS) [12] is a validated assessment tool measuring how well patient engagement is conducted. The Quality of Patient-Centered Outcomes Research Partnerships Instrument [13] is another assessment tool focusing on the meaningfulness of the patient partner role. The Public and Patient Engagement Evaluation Tool (PPEET) [14] assesses the quality of the engagement experience in which people were involved. In addition to these measures of the engagement process or experience, various measures exist to measure the impact of collaboration, often from a team science approach [15]. Yet, these measures focus largely on interpersonal dynamics, rather than the broader and often concrete impacts of engagement. No measures to date assess the impact that the engagement contributions have on the research itself, across the research lifecycle. This study aimed to address that gap.

### 1.1 Objective

This study aimed to develop and pilot test an assessment tool measuring the impact that lived/living experience engagement has on the research, called the *Measure of Engagement Tool for Research and lived Experience* (METRE).

## 2 Method

Using a sequential mixed-methods design, we began with a qualitative descriptive study of the impacts that engagement has on research, leading to the development, pilot testing, and refinement of an assessment tool. The project was underpinned by a scoping review of the literature on the impact of engagement and evidence gaps [1, 11]. The mixed-methods framework was selected in order to develop and pilot test the quantitative tool based on established qualitative data. The qualitative descriptive study informed the development of the tool, which was then pilot tested using qualitative and quantitative methods, which were interpreted together. A Lived/Living Experience and Caregiver Working Group was involved throughout the project, as described in Table 1. Table 1 is drawn from a recent reporting framework for lived experience engagement in research and was completed together with the Working Group members [16]. The findings are reported upon using the Good Reporting of A Mixed Methods Study (GRAMMS) checklist (Appendix A) [17].

**Table 1.** Reporting on lived/living experience and caregiver engagement in the research project.

|  |  |
| --- | --- |
| **Who** was involved? | We engaged six people with lived/living and caregiving experience at the onset of the project, which reduced to 3 members due to competing priorities. We added two new members, for a total of 5 people with lived/living and caregiving experience engaged throughout the bulk of the project. There was diversity in age range, but all were women. Several members had pre-existing relationships, which facilitated the development of positive working relations. |
| **What** were the activities, roles, and responsibilities? | The members were involved in all stages of the project. Notably, they discussed the project direction as a whole, reviewed and edited participant-facing materials for the full project, edited the focus group guide, and reviewed and refined the codebook for the qualitative study. For the METRE, they co-developed the scale, pre-pilot tested it, provided input on the instructional video, reviewed the pilot test results and revised the METRE and instructions in detail, provided additional input to facilitate the conduct of the study, and co-authored the paper. Two members were collaborators on the grant application and commented on its content prior to submission. |
| **How** did you go about the engagement process? | We held virtual Working Group meetings, supplemented by some asynchronous work. Meeting times were discussed with all and chosen by consensus. In the meetings, we created time to build relationships, with ice-breakers and dedicated time for casual communication. There were more people with lived/living and caregiver experience in the Working Group than academic researchers, which enhanced their power. The decision-making process was collaborative. The research lead and staff led meaningful engagement through their knowledge of issues surrounding lived experience and stigma. Members were given flexibility and choices, with opportunities to contribute in ways that fit with their schedules and interests. Accommodations included offering breaks for meetings longer than an hour, holding meetings virtually, and moving the meetings onto their preferred virtual meeting platform. Members were compensated promptly for their time. |
| **When** did engagement occur? | We held regular Working Group meetings ranging from 30 minutes (one meeting) to two hours. Meetings ranged from weekly to monthly, plus asynchronous time. There were a total of 27 |
|  | working group hours plus 8.5 hours of asynchronous work dedicated to this project. Engagement began in 2024 at the qualitative stage, continued to article submission in April 2026, and will continue thereafter for psychometric validation. The members report that this was sufficient time for ample engagement and it felt like an unrushed process. |
| **What** was the impact? | Engagement impacted the entire project. Working Group members report that the engagement provided opportunities to build their research capacity and their CVs. They explain that it felt like an inclusive process. For the academic researcher and research staff, it helped ensure that lived experience was represented in the project and provided opportunities for meaningful discussion rooted in real-world engagement experiences. All members of the Working Group, including people with lived/living experience, caregivers, and academic researcher/staff, learned about engagement through the process and expanded their knowledge. We conducted an informal evaluation of the engagement process, which helped understand areas of strength and areas of improvement. The high quality of the engagement that went into the early stages of the project helped the grant submission be funded at the first submission to the federal funder. |

### 2.1 Stage 1. Qualitative descriptive study

#### 2.1.1 Participants & recruitment

We recruited 28 lived/living experience and caregiver partners and 12 academic researchers with engagement experience for focus groups and individual interviews respectively. Participants had to be people with lived/living experience, caregivers, or academic researchers aged 16 or over with experience in research engagement settings in mental health and substance use health research, residing in Canada. Potential participants were contacted from previous Canada-wide studies on engagement (with consent and ethics approval) and by circulating a study flyer in our engagement networks.

#### 2.1.2 Procedure

Potential participants reached out to the research analyst for information, screening, and informed consent. Electronic informed consent was collected, guided by an e-consent framework on REDCap survey software [18]. Using a co-designed semi-structured interview guide, we completed five virtual focus groups and 12 individual interviews to understand participant perspectives on how engagement impacts the research. Focus groups and interviews took place on WebEx teleconferencing software, and were audio recorded, transcribed, and uploaded to NVivo for analysis. Participants received a $50 e-gift card in compensation for their time. Data collection took place between October 2024 and February 2025. Research Ethics Board approval was obtained from the Centre for Addiction and Mental Health.

#### 2.1.3 Measures

The demographic form was developed with feedback from the Lived/Living Experience and Caregiver Working Group to capture basic participant characteristics. The interview guide was co-designed based on our review of the literature and the perspectives of our Working Group. It focused on the areas in which lived/living experience and caregiver partners have impacts on a research project and the size of the impact. The interview guide is provided in Appendix B.

#### 2.1.4 Analysis

We used a framework analysis approach [19]. A codebook was developed to code the data into key areas of impact identified in the transcripts and based on the literature, including 1) research priority setting and developing research question, 2) grant applications, 3) concrete study planning, 4) recruitment processes, 5) data analysis and results interpretation, 6) knowledge translation and reporting, 7) the research climate, and 8) research quality. For each domain, people with lived/living experience, caregivers’, and academic researchers’ direct experiences were coded as small, medium and large engagement impacts. Through the coding process, we iteratively and inductively refined the codebook, dividing out some items into separate codes, for a final list of ten domains, and replacing ‘research quality’ with ‘research relevance.’ Qualitative results are presented briefly below, with representative quotes, with a focus on the scale development.

#### 2.1.5 Positionality

The research analyst (JH) is a full-time research analyst at the Centre for Addiction and Mental Health, with a master’s degree in psychology. She is new to the lived/living experience engagement research and is willing to learn from team members and participants. The scientific lead (LDH) is a scientist with an educational background in psychology, a research focus on lived/living experience engagement, and both qualitative and quantitative research skills. They were supported by the METRE Lived/Living Experience and Caregiver Working Group. The work was conducted out of a tertiary care mental health and addictions teaching and research hospital. While all members believe in the potential positive impacts of engagement, most have both highly positive and less positive impacts and all remained open to the breadth of participant responses.

### 2.2 Stage 2. Assessment tool development

The results of the qualitative descriptive study were used to guide the iterative development of the assessment tool. Specifically, 10 items were established, representing the themes in the qualitative data. Within those items, descriptive text was developed to describe no impact, a small impact, a medium impact, and a large impact, based on the sum of the dataset and substantial team discussions to illustrate general levels of engagement with consistency across items. Items, anchors, wording, and structure were iteratively reviewed and refined with the Lived/Living Experience and Caregiver Working Group and in consultation with a committee of academic researchers. The assessment tool was named the *Measure of Engagement Tool for Research and lived Experience* (METRE).

### 2.3 Stage 3. Pilot testing

The pilot study aimed to pilot test the METRE, with a view to refining it for the purposes of future validation. We selected a mixed-method framework for this stage in order to collect qualitative data from cognitive interviewing on acceptability and face validity, while also collecting participants’ quantitative responses to the METRE to explore the performance of the items.

#### 2.3.1 Participants & recruitment

New participants were recruited. Participants were 13 lived/living experience and caregiver partners and 10 academic researchers with research engagement experience, across mental health/substance use research and physical health research. Participants had to be aged 16 or over, residing in Canada, and self-report being from one of the participant groups of interest. They were recruited via an existing database of people with lived/living experience, caregivers, and academic researchers who had consented to be contacted about future research and from responses to a flyer circulated in the patient-oriented research community, such as Canadian provincial engagement support units.

#### 2.3.2 Procedure

A research analyst met with each participant to obtain informed consent and collect data. First, participants provided demographic data on the REDCap system [18]. They viewed an instructional video about the METRE and accessed the METRE and written instructions on the WebEx teleconferencing system, individually in the company of the research analyst. Participants were instructed to identify one project that they had worked on in using engagement and to complete the METRE with reference to that project. They went through a ‘think aloud’ cognitive interview process from the instructions to the completion of the METRE [20]. The research analyst probed the participants through the think aloud process if they were not spontaneously sharing their thoughts. Eight standard qualitative interview questions were then asked at the conclusion of the interview. The full interview was audio recorded. Recordings were transcribed automatically by the Webex system and transcripts were thoroughly cleaned by study staff members. Data collection took place between October 2025 and January 2026.

#### 2.3.3 Measures

The demographic information form consisted of basic information about the participant. The METRE is as described in Stage 3 above. We added one quantitative scaling question at the end of the METRE: “*As a whole, on a scale of 1 to 10, how much do you think engagement impacted the project?*” This was to help us understand impact in relation to the METRE total score and will be required for the full psychometric testing stage. The eight qualitative interview questions that were co-developed with the Working Group members addressed aspects of completing the METRE such as how well they understood various aspects of the METRE and instructions, what would have helped them understand it better, how well the ratings represented their experience of engagement, and how useful they think the METRE would be to support engagement. The interview guide is provided in Appendix C.

#### 2.3.4 Analyses

Demographic data were analyzed descriptively. METRE data were also analyzed descriptively, with attention to the mean, standard deviation, range, interquartile range, and distribution. The correlation between the quantitative scaling question and the METRE total score was analyzed using a Spearman correlation due to a small sample size (one missing, n = 22). Quantitative analyses of the pilot test data were compiled using SPSS 27 software. Item-level descriptive data are reported. Skewness and kurtosis were calculated for each item, with z-scores for significance testing [21]. Significance was examined at a conservative α <.01 given the small sample size. Think-aloud transcripts and qualitative interviews were uploaded into NVivo and analyzed using conventional content analysis [22]. Analyses were both inductive, with openness to innovative codes, and deductive, with a focus on identifying refinements to the scale. Missing data were minimal and thus handled using available-case analysis.

### 2.4 Stage 4. Post-pilot scale refinement

Upon completion of the pilot test and data analyses, the qualitative and quantitative results were brought together to the Lived/Living Experience and Caregiver Working Group for conjoint interpretation and to propose final refinements to the METRE. These were shared with team scientists for further review, whose comments were brought back to the Working Group for final decisions.

## 3 Results

### 3.1 Stage 1. Qualitative descriptive study

Participant characteristics for the qualitative descriptive study are provided in Table 2. There was some diversity in the sample across age, gender, ethnic background, and country of birth. Participants had experience with research from a lifespan development perspective, including child and youth research, adult research, and some geriatrics research.

**Table 2.**
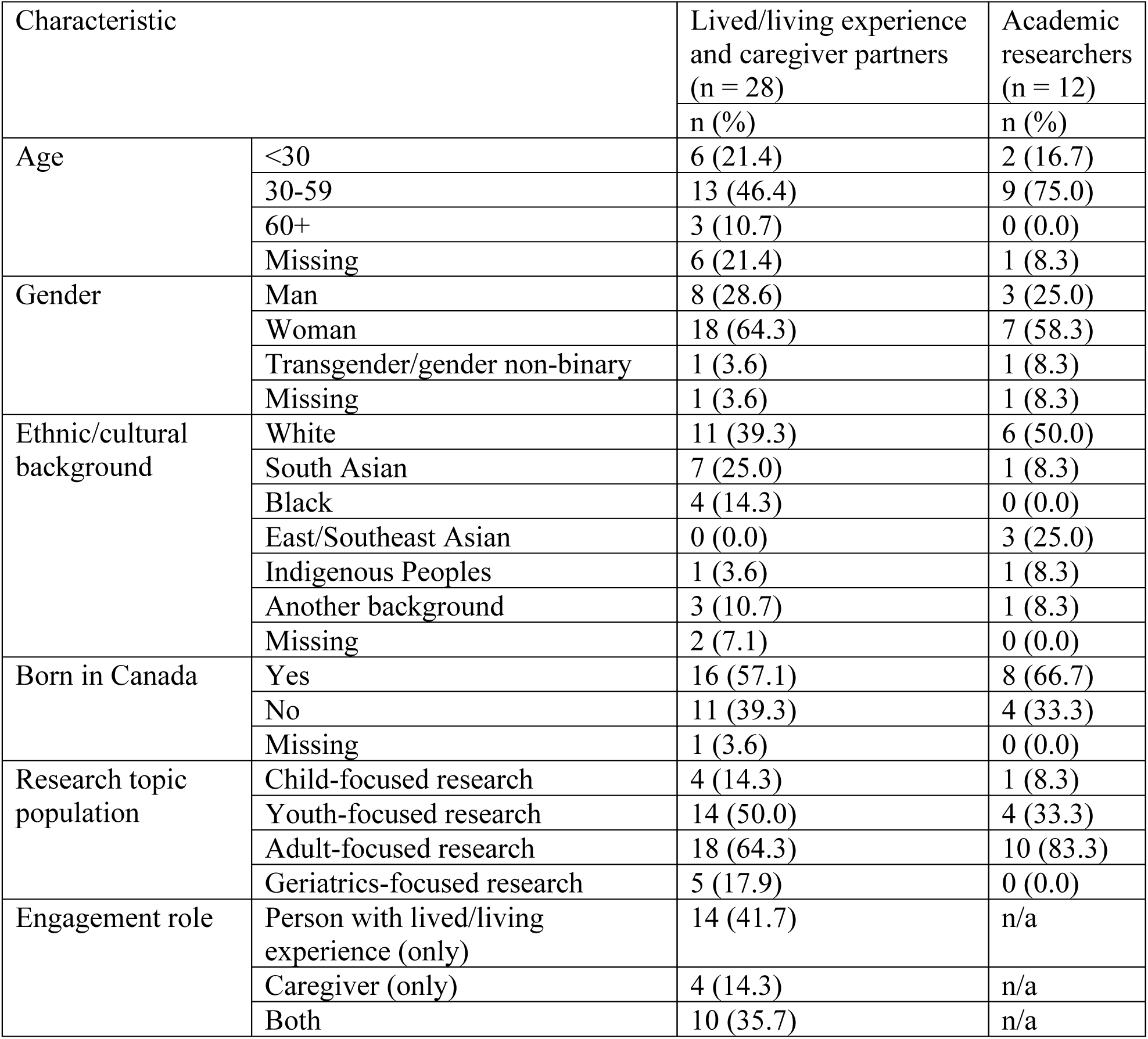
Demographic characteristics of the qualitative sample (N = 40)

The 10 domains of the METRE were derived from the qualitative findings briefly described below. Quotes represent the range of impacts that lived experience and caregiver engagement has on the research as described by participants.

1. Research priority setting. Some participants had no experience with research priority setting: “*So as far as I can remember, nobody was involved at the like conceptualization stage.” [Participant #1, Lived/living experience/caregiver participant]* Others had some small impact on the research priority setting stage, such as “*point[ing] out some additional ways that we could use the data that we’re trying to compile*.”*[Participant #2, Lived/living experience/caregiver participant]* However, some participants reported a more moderate contribution to this stage of the research, such as *“reviewing funding proposals on dementia and depression” [Participant #3, Lived/living experience/caregiver participant],* where they were valued and were invited to provide their opinions about the research. Some participants surpassed this moderate level, reporting large impacts on research priority setting: *“they influenced greatly what the education the learning objectives were, and how the curriculum, the content and how the curriculum was gonna be delivered*.” *[Participant #4, Academic researcher]*
2. Developing research questions. There was an interest, but sometimes a lack of involvement, in in the early stages of the research: “*I’d like to be involved with earlier aspects, like things like deciding which research questions we want to study or even which grants we want to apply to*.” *[Participant #5, Lived/living experience/caregiver participant]* Some participants reported some minimal or light involvement in the question development stage: “[*B*]*ecause I was not there at the beginning stage of the brainstorming, I felt like I couldn’t really change the subject of what we wanted to discuss, but sort of the course of that*.” *[Participant #2, Lived/living experience/caregiver participant]* Others described being able to moderately contribute to sub-questions more thoroughly, after the initial questions had been set: “*there is that main research question, but under that you could have like subcategories of focus areas, right*?” *[Participant #6, Lived/living experience/caregiver participant]* Lastly, a few participants described large impacts of people with lived/living experience on the research question setting stage: “*These youth come to us with mental health lived experiences, and they’re telling us this, this, this, this. So, they inform us, and then we, you know, use our research methodologies just to refine it (…) but it’s the intention remains the same*.” *[Participant #7, Academic researcher]*
3. Grant applications. Not all participants were engaged at the grant application stage: “*Patients were engaged after we got funding*.” *[Participant #4, Academic researcher]* However, sometimes they were engaged in grant applications with a small impact: “*I reviewed the grant application, I shared my insights and feedback about that. I provided my biosketch*.” *[Participant #8, Lived/living experience/caregiver participant]* This extended to potentially slightly stronger impacts, with involvement at the start of the grant application stage: “*When we started writing of the proposal, when we prepared the proposal, we kind of consulted them and take some insights from them*.” *[Participant #9, Academic researcher]* The largest contributions were seen through participant quotes such as the following: “*For example, at least one principal investigator should be a person with lived experience, and having that outlined very concretely in a grant eligibility or whatever.*” *[Participant #10, Academic researcher]*
4. Concrete study planning. The concrete study planning process, including the REB process, was often not an area in which people with lived/living experience and caregivers were engaged “*I don’t think I’ve been involved in a research project where—until they’re past ethics*.” *[Participant #11, Lived/living experience/caregiver participant]* However, some acknowledgement of being involved in the study planning was observed: “*I’ve more so been involved with the actual process of creating the study.*” *[Participant #5, Lived/living experience/caregiver participant]* Medium impacts at this stage were also indicated by some participants: *We had conversations with the whole team with lived expert folks of like what should this look like? And then we went away and drafted that and then brought it back.*” *[Participant #12, Academic researcher]* Sometimes a more thorough co-design approach was indicated: “*So, I helped to redo their photovoice sessions. I redid their photovoice manuals, their user manuals, and their participant manuals.*” *[Participant #11, Lived/living experience/caregiver participant]*
5. Recruitment processes. With regard to the recruitment process, some participants expressed that this was an early study stage in which they were not involved: “*As a family member, I was not involved in these early stages of creating the studies and recruitment*.” *[Participant #3, Lived/living experience/caregiver participant]* Sometimes lived/living experience and caregiver partners contributed in small ways to recruitment through a naturally occurring process: “*So I think they inadvertently helpfully helped with like word of mouth recruitment, but they were not tasked to do that if that makes sense.*” *[Participant #13, Academic researcher]* A more intentional, moderate level of impact was seen when recruitment challenges were encountered: “*I helped with figuring out some innovative approaches to capturing some possible participants within the community.*” *[Participant #6, Lived/living experience/caregiver participant]* More substantial engagement, including co-design, was also seen at this stage: “*So like creating like youth-friendly recruitment materials and things like that. That’s something where I’ve seen a lot of like co-design happen.*” *[Participant #14, Academic researcher]*
6. Data analysis. Often, there was no engagement at the data analysis level: “*No, ‘cause the data analysis was planned a priori. So, they did not have any input into that.*” *[Participant #15, Academic researcher]* Some had an impact ranging from small to large, notably in terms of qualitative data analysis: “*I volunteered to take on a lot of the analysis work. And because it was like a peer position, I still had to sort of look over the codes that other people had done.*” *[Participant #2, Lived/living experience/caregiver participant]* Lastly, a high degree of engagement is observed through one participant’s experience: “*We were taught how to pick out themes, analyze the data, transcribe the data, use grounded theory methodology to process the information*.” *[Participant #16, Lived/living experience/caregiver participant]* However, this was largely limited to qualitative data analysis.
7. Results interpretation. The interpretation of results was an area in which engagement was common; indeed, nobody reported no impact in this area. Small impacts were indicated by some participants, in the form of “confirmation”. “*It was like, does this like resonate with your experience that you’ve had*?” *[Participant #1, Academic researcher]* Moderate involvement in interpretation was also observed: “*I read over the theme summary document, and I provided, like it was in a Google Docs sheet, so I just like put in my comments. And they asked for feedback, and met with them a couple of times, so I just in those meetings went over the feedback that I’ve written out.*” *[Participant #1, Lived/living experience/caregiver participant*] In some cases, more substantial engagement was reported, where participants offered novel interpretations that were relevant but that the academic researchers had missed: “*[W]e asked them, do they think older people would be more or less sensitive to cannabis? And we had hypothesized they’d be more sensitive because of —their metabolism is slower, so the cannabis would last longer. But they came up with the opposite. They said, well, no, people who are older would have used cannabis longer for many more years, so they would be more tolerant. And I didn’t like that idea when they first proposed it. And I didn’t think that was very, you know, based like on the data but as it turns out, they were, I think they were right*.”*[Participant #15, Academic researcher]*
8. Knowledge translation and reporting. Some participants did not make a substantial contribution to the knowledge translation and reporting stage, despite a wish to do so: “*No, I don’t have experience in that area, but that would be amazing*.” *[Participant #13, Academic researcher]* Some small impacts were reported: “*It was more like small design related.*” *[Participant #17, Lived/living experience/caregiver participant]* More moderate contributions are described by an academic researcher participant: “*So, I usually do most of the writing and then I ask people with lived experience, well, other researchers, including people with lived experience, on my team, to complete certain sections*.” *[Participant #18, Academic researcher]* However, there were many examples of large contributions to the knowledge translation stage, as this is a stage in which lived/living experience and caregiver partners are often heavily engaged: “*They get to decide everything with respect to how knowledge gets translated. You know, my role here is made basically to ask questions to guide them back to, (…) these are our research questions, this is what we’re trying to answer.” [Participant #19, Academic researcher]*
9. The research climate. The research climate was not always positively impacted by engagement: “*So it feels sometimes it’s like ok yes, I understand this is the box. (…) [T]his is what the container is for me to participate*.”*[Participant #11, Lived/living experience/caregiver participant]* Small to medium impacts were also observed: “*[Y]ou’re treated really nicely, your opinions are valued, and it’s very positive environment when you’re actually doing the work*.” *[Participant #20, Lived/living experience/caregiver participant]* Large impacts were described by several academic researchers, including “*I think it goes without saying like there’s this reciprocal nature of partnership. So, while I’ve talked a little bit about the young people gaining confidence and skills in a number of ways, like certainly I have learned a ton from, you know, not only the lived experiences, but all of the skills and like the multiple facets of people’s identities and that they bring into research*.” *[Participant #21, Academic researcher]*
10. Research relevance. In terms of research relevance, few participants mentioned that there was no impact or a small impact of engagement. Medium and large impacts were far more common, with a medium impact described by a lived/living experience participant: “*I think people with lived experience can look at that and say, ‘yeah, that’s a valid point,’ or ‘yeah that’s that could be good to understand that,’ because you can base from your own experience, how relevant that particular topic is.*” *[Participant #22, Lived/living experience/caregiver participant]* Large impacts were described by an academic researcher: “*more relevant study findings, the more nuanced interpretation of the data, and then the development of really youth-friendly outputs that help to demonstrate our results to the broader community, and will hopefully help with uptake and reach*.” *[Participant #21, Academic researcher]*

### 3.2 Stage 2. Measure development

After analyses of the qualitative descriptive data, the pilot version of the METRE was progressively generated and refined with the Lived/Living Experience and Caregiver Working Group and team scientists. It consisted of ten items, each with a four-point impact scale. The ten items submitted to pilot testing were 1) research priority setting, 2) developing research question, 3) grant applications, 4) concrete study planning, 5) recruitment processes, 6) data analysis, 7) results interpretation, 8) knowledge translation and reporting, 9) the research climate, and 10) research relevance. The rating scale included ‘no impact,’ a ‘small positive impact,’ a ‘medium positive impact,’ and a ‘large positive impact.’ One open-ended field was provided at the end of the scale to identify any negative impacts. Each level of impact was briefly described in table format to help respondents select a score. We also co-developed print instructions and an instructional video to help respondents understand the METRE and the scoring process.

### 3.3 Stage 3. Pilot testing

Participant characteristics for the pilot study are provided in Table 3. While the sample was small, there was some diversity across demographic characteristics and participants had experience in research across the lifecycle.

**Table 3.** Demographic characteristics of the pilot study sample (N = 23)

| Characteristic |  | Lived/living experience and caregiver partners (n = 13) | Academic researchers (n = 10) |
| --- | --- | --- | --- |
|  |  | n (%) | n (%) |
| Age | <30 | 5 (38.5) | 5 (50.0) |
|  | 30-59 | 6 (46.2) | 5 (50.0) |
|  | 60+ | 2 (15.4) | 0 (0.0) |
| Gender | Man | 4 (30.8) | 3 (30.0) |
|  | Woman | 7 (53.8) | 7 (70.0) |
|  | Transgender/gender non-binary | 2 (15.4) | 0 (0.0) |
| Ethnic/cultural background | White | 6 (46.2) | 5 (50.0) |
|  | South Asian | 4 (30.8) | 1 (10.0) |
|  | East/Southeast Asian | 2 (15.4) | 1 (10.0) |
|  | Black | 1 (7.7) | 0 (0.0) |
|  | Another background | 0 (0.0) | 2 (20.0) |
|  | Multiple backgrounds | 0 (0.0) | 1 (10.0) |
| Born in Canada | Yes | 7 (53.8) | 4 (40.0) |
|  | No | 6 (46.2) | 5 (50.0) |
|  | Missing | 0 (0.0) | 1 (10.0) |
| Research topic population | Child-focused research | 5 (38.5) | 4 (4.0) |
|  | Youth-focused research | 8 (61.5) | 7 (7.0) |
|  | Adult-focused research | 12 (92.3) | 8 (8.0) |
|  | Geriatrics-focused research | 5 (38.5) | 3 (3.0) |
| Engagement role | Person with lived/living experience (only) | 6 (46.2) | n/a |
|  | Caregiver (only) | 1 (7.7) | n/a |
|  | Both person with lived/living experience and caregiver | 6 (46.2) | n/a |

#### 3.3.1 Qualitative feedback

Participants perception of the METRE was positive overall, although they made specific suggestions for refinement. Our findings are organized into three main themes: 1) The METRE is easy and straightforward to understand and complete, 2) The METRE provides a thorough evaluation of the impact of engagement, and 3) Some areas of improvement are recommended. Each theme is described by subthemes (see Table 4 for details).

**Table 4.** Themes and subthemes generated from the think-aloud pilot test stage of the project.

| Themes | Subthemes |
| --- | --- |
| 1. The METRE is easy and straightforward to understand and complete. | 1a. Instructions and video are helpful. |
|  | 1b. Items are easy to understand and align with their experience of engagement. |
| 2. The METRE provides a thorough evaluation of the impact of engagement. | 2a. Covers the full research process. |
|  | 2b. Relevant evaluation of engagement. |
|  | 2c. Will set the stage for good engagement practices. |
| 3. Some areas of improvement are recommended. | 3a. Some areas for improvement in instructions and layout suggested. |
|  | 3b. Some areas for item improvement suggested. |
|  | 3c. Sometimes difficulty discriminating between levels. |

##### Theme 1. The METRE is easy and straightforward to understand and complete

Participants generally described the METRE as easy and straightforward to understand and complete. The instructional support was a significant factor in usability. The provided instructions (e.g., general instructions, sample project) and introductory video were reported to be helpful for most participants. Many participants also thought that the METRE’s ten items were easy to understand and matched their experience being engaged in research.

> *“I liked it was very easy to fill out. It was very easy to understand, and the way you have it scaled is nice.” [Participant #1, Lived/living experience/caregiver participant]*

##### Theme 2. The METRE provides a strong evaluation of the impact of engagement

Participants generally described METRE as a meaningful and relevant tool to assess the impact that engagement has on research. This extended to people with lived/living experience, caregivers, and academic researcher participants. Many noted that the tool captured engagement across the full research process, including early-, mid-, and later-stage activities and overall perceptions. Participants also reported that the content of METRE reflected their experiences of engagement research and provided a relevant evaluation of how lived/living experience and caregiver partner involvement was integrated into research projects. Some participants also mentioned that completing METRE prompted reflection on engagement practices and had the potential to set the stage for future research activities.

> *“I do think it would be quite useful, because there really isn’t much existing. […] I think it sets a high standard, which is a good thing. Because even like looking back, having done the rating for our project, it gave me pause to see some areas where we didn’t have as much impact.” [Participant #2, Academic researcher]*

##### Theme 3. Some areas of improvement are recommended

Although overall feedback was positive, participants identified several areas for improvement. Suggestions related to instructions and layout included simplifying content, providing clearer guidance, and rearranging the order of the instruction sections to further enhance usability. Participants also identified specific items that could benefit from refinement, such as clearer wording, additional plain-language definitions, additional areas of impact, or examples to improve alignment with diverse research contexts. In addition, some participants reported difficulty deciding on response options for certain items, including challenges distinguishing between response levels. These suggestions informed the refinements to improve scale clarity and response consistency.

> *“I think just with the statements, as I was saying, like if they can be a little bit more clear. I understood what to do and stuff, but I think some of them were a little bit(…) like I had to read through every item [levels of impact] and when I would read the large impact, then that’s where more of the description would be.” [Participant #3, Lived/living experience/caregiver participant]*

#### 3.3.2 Quantitative findings

Quantitative results on the METRE pilot test are presented in Table 5. Items showed variability in responses, with responses ranging from 1 (no impact) to 4 (large positive impact) for all except one item, which had no endorsements of the ‘no impact’ anchor. Medians ranged from 2 to 4. None of the skewness or kurtosis values were statistically significant at the conservative <.01 level. These are acceptable properties of the pilot test items.

**Table 5.** Item-level quantitative results for the pilot test data.

| Item number | Mean | SD | Range | Median [IQR] | Skewness (SE) | z | Kurtosis (SE) | z |
| --- | --- | --- | --- | --- | --- | --- | --- | --- |
| Item 1 | 2.61 | 0.99 | 1-4 | 3 [2-3] | -0.02 (0.48) | -0.04 | -0.94 (0.94) | -1.01 |
| Item 2 | 2.70 | 1.06 | 1-4 | 3 [2-4] | -0.32 (0.48) | -0.66 | -1.01 (0.94) | -1.11 |
| Item 3 | 2.26 | 1.14 | 1-4 | 2 [1-3] | 0.45 (0.48) | 0.94 | -1.16 (0.94) | -1.24 |
| Item 4 | 3.04 | 0.98 | 1-4 | 3 [2-4] | -0.74 (0.48) | -1.53 | -0.35 (0.94) | -0.38 |
| Item 5 | 3.04 | 1.19 | 1-4 | 4 [2-4] | -0.81 (0.48) | -1.68 | -0.94 (0.94) | -1.01 |
| Item 6 | 2.61 | 1.16 | 1-4 | 3 [2-4] | -0.10 (0.48) | -0.20 | -1.44 (0.94) | -1.54 |
| Item 7<sup>1</sup> | 2.96 | 1.07 | 1-4 | 3 [2-4] | -0.65 (0.48) | -1.35 | -0.77 (0.94) | -0.82 |
| Item 8<sup>1</sup> | 3.05 | 1.05 | 1-4 | 3 [2-4] | -0.65 (0.49) | -1.32 | -0.87 (0.95) | -0.91 |
| Item 9<sup>1</sup> | 3.27 | 0.88 | 1-4 | 3.5 [3-4] | -1.05 (0.49) | -2.15 | 0.45 (0.95) | 0.47 |
| Item 10<sup>1</sup> | 3.32 | 0.78 | 2-4 | 3.5 [3-4] | -0.65 (0.49) | -1.33 | -0.99 (0.95) | -1.04 |

SD = standard deviation; IQR = interquartile range; SE = standard error; <sup>1</sup>Items 7 through 10 are based on n = 22 due to missing data for one participant.

The quantitative scaling question, where participants were asked to quantify the degree of impact that engagement had on their project on a 10-point scale, was moderately correlated with average scores on the METRE. The Spearman correlation was *r*_s_ = .42, (*p*=.053, 95% CI = [-.02, .72]), suggesting a moderate association.

### 3.4. Stage 4. Post-pilot scale refinement

All items were revised through multiple meetings and asynchronous work with the Working Group, by interpreting both the qualitative and quantitative findings in a mixed-methods framework. Item names and descriptions were iteratively refined for greater clarity and to ensure applicability across the broadest possible spectrum of research projects, including projects not collecting data from human subjects and those not funded by grant applications. The quantitative data helped contextualize the qualitative data and provided confidence regarding item distributions. We also sent the METRE to a copy-editing service for final adjustments and a quality check. The final METRE consists of 10 items, which are rated a response scale of zero to three points, ranging from no impact to a large positive impact. The final areas of impact are 1) research topic and priority setting, 2) developing research questions, 3) finding funding, 4) project planning, 5) participant recruitment and data collection, 6) data analysis and results generation, 7) results interpretation, 8) sharing results and reporting, 9) research climate, 10) research relevance. The final METRE retained for the psychometric testing stage is provided in Appendix D.

## 4. Discussion

While lived/living experience and caregiver engagement is rapidly growing in popularity among contemporary research teams, the assessment of engagement is in its early stages [11]. This study aimed to develop a novel measurement tool to measure the impact that engagement has on research. A qualitative phase provided areas of impact to measure, leading to the development of the METRE. Pilot testing provided refinements to the assessment tool. The resulting tool consists of 10 domains of perceived impact in which lived/living experience and caregiver engagement may affect research to varying degrees.

This METRE was designed to assess the perceived impact of engagement on research with both qualitative and quantitative insights, to be completed together among people with lived/living experience, caregivers, and academic researchers, directly addressing an established evidence gap[11]. The quantitative scores can be used to evaluate levels of impact across the research process for different types of engagement models and to guide research teams toward engagement models with a stronger impact. The item descriptions can be used to inform the team in more detail about the areas of strength and of untapped potential. We recommend that it be completed in conjunction with a robust reporting of the methods of engagement in the research project [16, 23]. It is important to note that the METRE is a measure of *perceived* impacts of engagement on research, which is the case for the bulk of the impact literature to date [1]. Objective evaluations of impact might include thorough reviews of meeting notes, action items, and project decisions in relation to the final project deliverables, or even trials of the feasibility of research designs developed through engagement versus without.

It should also be noted that in the qualitative descriptive stage, the ‘high impact’ level was described most often by academic researchers, rather than people with lived/living experience and caregivers. This might reflect a response or experiential bias in the scale, i.e., it may be that academic researchers perceive the impacts of engagement to be larger. It is also possible that they have more of an overarching view of the research or are more experience with engaged research, which might shift their descriptions to a higher impact level compared to the perceptions of people with lived/living experience and caregivers. This should be further examined in the psychometric testing stage.

Pilot data show that all stages of the research process were impacted by engagement, which reflects the nascent literature on impact [1]. This shows that patient-oriented research teams are working together in positive and productive ways. However, we identified that data analysis was the project stage with the least engagement, at least among pilot test participants. This has been identified as a gap in the past [2, 24]. However, people with lived/living experience and caregivers can be engaged in data analysis, and this therefore appears to be an area for further development in engagement spaces.

The next step in this line of research is to determine the psychometric properties of the METRE. Its construct validity, reliability, and factor structure have yet to be established. An initial validation project is currently under way in an international sample of lived/living experience and caregiver partners and academic researchers across domains of health research. It should be noted that there may yet be changes to the METRE as this process is pursued, including item reduction and refinement. Once that next important stage is complete, the tool will be disseminated and placed in the public domain to encourage widespread use. Pending validation, research teams are encouraged to use it and interpret its findings with caution. Other recent work has also begun exploring new approaches to documenting and evaluating the impact of lived/living experience and caregiver engagement and co-production in research [25]. These various initiatives should be monitored as the measurement science of engagement continues to grow.

Despite the importance of developing new measures to support measurement science in engagement, it is also important to remember that engagement is called for as an ethical imperative [26]. Indeed, engagement is a democratizing process and a matter of epistemic justice for lived/living experience populations that has intrinsic value [26]. Therefore, even if engagement is observed to not have the positive impacts that are sought, it is still a valid and important activity to for researchers to undertake [27, 28]. Negative assessments of impact should never undermine the importance of lived/living experience and caregiver engagement for the healthcare system.

### 4.1 Strengths and limitations

This study was co-produced through extensive collaboration with the Lived/Living Experience and Caregiver Working group, which is a project strength. The paper systematically illustrates the full spectrum of the research process, from the underlying qualitative stage to the end of the pilot test. A proportion of participants were youth under age 30, and over half of participants had youth-oriented research experience, making the findings relevant to research from a lifespan development perspective. However, limitations must be kept in mind when interpreting the findings. The qualitative study was focused on people with lived/living experience of the mental health/substance use health spectrum, which may differ from those engaging in physical health research. Completing the think aloud process may have influenced the scores provided due to response bias, which was indicated by one participant. The final psychometric testing stage will not have this bias, since there will be no think-aloud process. Given the small sample size, people with some demographic characteristics might have been missed. Notably, the majority of participants were women. The Lived/Living Experience and Caregiver Working showed the same bias, as all members were women. A further limitation is with regard to selection bias: it is possible that people with more experience of high-impact and positive engagement and in institutions with strong support for engagement were more interested in participating in the study. Future research on a larger and more diverse sample is required to address these limitations.

## 5 Conclusions

The METRE is a new, co-developed assessment tool designed for completion by lived/living experience and caregiver partners and academic researchers across the domains of health research. It has been developed together with people with lived/living experience and caregivers to provide relevant assessments of the subjective impact of engagement on research projects.

Still pending is the evaluation of the psychometric properties of the tool scores across different research settings. Nevertheless, this tool adds to the limited set of assessment tools available to measure aspects of lived/living experience and caregiver engagement in research.

## Supporting information

Diagram

## Data Availability

The data that support the findings of this study are available on request from the corresponding author with approval from the Research Ethics Board of the Centre for Addiction and Mental Health. The data are not publicly available due to privacy or ethical restrictions.

## Statements and Declarations

### Availability of data and materials

The data that support the findings of this study are available on request from the corresponding author. The data are not publicly available due to privacy or ethical restrictions.

### Ethics approval and consent to participate

Research ethics board approval was received from the Centre for Addiction and Mental Health, REB# 2024-070, 2025-064, 2025-177. All participants provided electronic signed informed consent.

### Authors’ contributions

**Lisa D. Hawke**: conceptualization, methodology, validation, data curation, formal analysis, visualization, writing – original draft, writing – review & editing, supervision, project administration, funding acquisition. **Jingyi Hou**: Conceptualization, methodology, validation, formal analysis, resources, writing – original draft, writing – review & editing. **Katie Upham**: conceptualization, methodology, validation, writing – review& editing**, Mary Rose van Kesteren**: conceptualization, methodology, validation, writing – review& editing**, Charlotte Munro**: conceptualization, methodology, validation, writing – review& editing, **Shoshana Hauer**: conceptualization, methodology, validation, writing – review& editing**, Claudia Sendanyoye**: conceptualization, methodology, validation, writing – review& editing. **Tanya Halsall:** conceptualization, methodology, validation, writing – review& editing**, Lena C. Quilty**: conceptualization, validation, writing – review& editing, **Clayon Hamilton:** conceptualization, methodology, validation, writing – review& editing, S**kye Barbic**: conceptualization, methodology, validation, writing – review& editing, **Wei Wang**: conceptualization, methodology, validation, writing – review& editing

## Acknowledgements.

We would like to thank Wuraola Dada-Phillips and Shelby McKee for the Stage 1 focus group and interview facilitation. We acknowledge the Ontario SPOR Support Unit, Alberta SPOR Support Unit, Maritime SPOR unit, BC SUPPORT Unit, Passerelle National Training Entity in Patient Engagement, the SPOR Evidence Alliance, and REACH BC assistance in participant recruitment. We further acknowledge that this work was conducted at CAMH, which is situated on lands that have been occupied by First Nations for millennia; lands rich in civilizations with knowledge of medicine, architecture, technology and extensive trade routes throughout the Americas. The site of CAMH appears in colonial records as the council grounds of the Mississaugas of the New Credit (as their name in 1860), today known as the Mississaugas of the Credit.

## Appendix A: Good Reporting of A Mixed Methods Study (GRAMMS)

| Criterion | Page |
| --- | --- |
| (1) Describe the justification for using a mixed methods approach to the research question | 3 |
| (2) Describe the design in terms of the purpose, priority and sequence of methods | 3 |
| (3) Describe each method in terms of sampling, data collection and analysis | 3-8 |
| (4) Describe where integration has occurred, how it has occurred and who has participated in it | 3, 13-15 |
| (5) Describe any limitation of one method associated with the present of the other method | 16, 17 |
| (6) Describe any insights gained from mixing or integrating methods | 15, 16 |

O'Cathain A, Murphy E, Nicholl J. The quality of mixed methods studies in health services research. J Health Serv Res Policy. 2008;13(2):92-98

## Appendix B. Stage 1 Qualitative Descriptive Study Semi-structured Interview Guide

1. First, think about a time when you were involved in a PWLE-engaged study. What did that look like?

a. How were PWLE engaged?
b. What are the stages of a project where PWLE have a big impact on the project?
c. What kinds of impacts did they have on the project?
2. How does PWLE engagement impact the concrete planning of the research project, like creating study and recruitment materials?

a. Have you ever been involved in this process? What changes were made?
b. What about developing project materials?
c. What were the challenges and successes?
d. POLL: How big of a change do you think you made?

i. 1 Very small
ii. 2 Small
iii. 3 Medium
iv. 4 Somewhat large
v. 5 Large
e. What would it look like if PWLE made a small contribution to this stage?
f. What would a medium contribution look like?
g. What would a large contribution look like?
3. How does PWLE engagement impact the research ethics board process?

a. Have you ever been involved in this process? What changes were made?
b. What were the challenges and successes?
c. POLL: How big of a change do you think you made?

i. 1 Very small
ii. 2 Small
iii. 3 Medium
iv. 4 Somewhat large
v. 5 Large
d. What would it look like if PWLE made a small contribution to this stage?
e. What would a medium contribution look like?
f. What would a large contribution look like?
4. How does PWLE engagement impact the participant recruitment process?

a. Have you ever been involved in this process? What changes were made?
b. What were the challenges and successes?
c. POLL: How big of a change do you think you made?

i. 1 Very small
ii. 2 Small
iii. 3 Medium
iv. 4 Somewhat large
v. 5 Large
d. What would it look like if PWLE made a small contribution to this stage?
e. What would a medium contribution look like?
f. What would a large contribution look like?
5. How does PWLE engagement impact the data analysis process?

a. Have you ever been involved in this process? What changes were made?
b. What were the challenges and successes?
c. POLL: How big of a change do you think you made?

i. 1 Very small
ii. 2 Small
iii. 3 Medium
iv. 4 Somewhat large
v. 5 Large
d. What would it look like if PWLE made a small contribution to this stage?
e. What would a medium contribution look like?
f. What would a large contribution look like?
6. How does PWLE engagement impact the interpretation process?

a. Have you ever been involved in this process? What changes were made?
b. What were the challenges and successes?
c. POLL: How big of a change do you think you made?

i. 1 Very small
ii. 2 Small
iii. 3 Medium
iv. 4 Somewhat large
v. 5 Large
d. What would it look like if PWLE made a small contribution to this stage?
e. What would a medium contribution look like?
f. What would a large contribution look like?
7. How does PWLE engagement impact the knowledge translation and reporting process?

a. Have you ever been involved in this process? What changes were made?
b. What were the challenges and successes?
c. POLL: How big of a change do you think you made?

i. 1 Very small
ii. 2 Small
iii. 3 Medium
iv. 4 Somewhat large
v. 5 Large
d. What would it look like if PWLE made a small contribution to this stage?
e. What would a medium contribution look like?
f. What would a large contribution look like?
8. How does PWLE engagement affect the choice of research priorities or determining research questions?

a. Have you ever been involved in this process? What changes were made?
b. What were the challenges and successes?
c. POLL: How big of a change do you think you made?

i. 1 Very small
ii. 2 Small
iii. 3 Medium
iv. 4 Somewhat large
v. 5 Large
d. What would it look like if PWLE made a small contribution to this stage?
e. What would a medium contribution look like?
f. What would a large contribution look like?
9. How does PWLE engagement impact the grant application process, when the initial research plan is developed?

a. Have you ever been involved in this process? What changes were made?
b. Are there any budget contributions?
c. What were the challenges and successes?
d. POLL: How big of a change do you think you made?

i. 1 Very small
ii. 2 Small
iii. 3 Medium
iv. 4 Somewhat large
v. 5 Large
e. What would it look like if PWLE made a small contribution to this stage?
f. What would a medium contribution look like?
g. What would a large contribution look like?
10. Now think about the research climate as a whole. How does engagement affect the research beyond these study processes?
11. What about relationships? The spirit of teamwork? The way the team reflects on the topic? The relevance of the research? The quality of the research? Other aspects?
12. What were the challenges and successes?
13. POLL: How big of a change do you think you made?

i. 1 Very small
ii. 2 Small
iii. 3 Medium
iv. 4 Somewhat large
v. 5 Large

a. What would it look like if PWLE made a small contribution to this?
b. What would a medium contribution look like?
c. What would a large contribution look like?
14. Are there any other ways that PWLE engagement impacts the research process?

## Appendix C. Stage 3. Pilot Testing Interview Guide

Qualitative questions

1. What was it like to complete the METRE?
2. How well do you feel you understood how to complete the METRE?

a. How well did the video prepare you?
b. How well did the instructions prepare you?
3. How well did you understand what the questions were asking?
4. What would have helped you understand it better?
5. How well does the METRE reflect the impacts of PLLEX-C on research?
6. How well did the rating scale reflect your perceptions of a small, medium, and large impact on the project?
7. How useful do you think the METRE would be to support engagement?
8. As a whole, on a scale of 1 to 10, how much do you think PLLEX-C engagement impacted the project?
9. Is there anything else you would like to tell us about the METRE or your experience completing it?

## Appendix D-1. METRE

### Measure of Engagement Tool for Research and lived Experience (METRE)

#### Instructions

People with lived and living experience and family/caregivers are increasingly engaged in research as advisors, collaborators, and co-researchers, beyond research participation roles. The METRE measures the impact of engagement, to help people with lived experience, caregivers, and researchers better understand the impact of engagement across the health research sector. By “impact,” we mean how much influence engagement had on the process and outcome of the research project.

To complete the METRE, you’ll rate 10 research areas where engagement may have an impact on the project. For each area, choose 0–3, where a zero indicates that engagement had no impact on the research and a three indicates a large positive impact. An open-ended question about any possible negative impacts is provided at the end.

Please think back to a specific research project and rate that project. When selecting a number, consider aspects like the following:

- when and how frequently people with lived/living experience and caregivers were engaged (e.g., early vs. later stages)
- the amount of input they provided
- the extent to which their input influenced decisions
- the extent to which their input changed the project for the better

If you are undecided between two responses, select the one that best reflects the overall pattern of impact. You can also clarify your response in the “Describe” section.

#### NOTE

A larger impact score does NOT mean the overall project is better. It reflects the size of the engagement’s positive impact on the research. Each project and team is unique.

An example of a completed METRE is provided on the next page.

##### Suggested citation

Lisa D. Hawke, Jingyi Hou, Katie Upham, Mary Rose Van Kesteren, Charlotte Munro, Shoshana Hauer, Claudia Sendanyoye, Tanya Halsall, Lena C. Quilty, Clayon Hamilton, Skye Barbic, Wei Wang. (2026). Measure of Engagement Tool for Research and lived Experience (METRE). Centre for Addiction and Mental Health, Toronto, Canada.

#### Sample Project

Below is a sample table illustrating a METRE that was completed by people with lived and living experience, together with a diagram illustrating the results.

*Note that this is not an actual project, but a sample for illustrative purposes*.

##### I am a

☒ Lived experience advisor, collaborator, co-researcher
☐ Family/caregiver advisor, collaborator, co-researcher
☐ Scientist or academic investigator with a research institution
☐ Research staff or academic trainee

###### Please use the scale below

0 No impact
1 Small positive impact
2 Medium positive impact
3 Large positive impact

| Area of Impact | Score | Describe |
| --- | --- | --- |
| 1. Research topic & priority setting | 0 | The team already had their priorities set when we joined the team. |
| 2. Developing research questions | 1 | While the main research questions were already established, we suggested improvements to some of the questions. |
| 3. Finding funding | 2 | We talked through the project at a few meetings and made suggestions for improvements, which were taken into account. |
| 4. Project planning | 3 | We worked together with the scientists to create all of the documents that participants see. We helped them choose the questionnaires and gave them quite a few recommendations. The scientists used our suggestions to improve the project. |
| 5. Participant recruitment & data collection | 2 | We helped develop the study flyer and emails. We had ideas about where the scientists could find study participants. We sent out recruitment flyers to find participants. |
| 6. Data analysis & results generation | 2 | We reviewed the data analyses and suggested some new analyses that could be done, which the team then carried out and presented to us. |
| 7. Results interpretation | 2 | We discussed how we understood the results, which was different from the way the scientists understood them. The scientists refined their results based on our perspectives. |
| 8. Sharing findings & reporting | 3 | We helped create social media ads presenting the findings and we shared the social media posts. We co-authored the paper and provided our perspectives throughout. |
| 9. Research climate | 2 | We developed a strong trusting relationship with the scientists. It was a safe space to work together as a team. |
| 10. Research relevance & applicability | 3 | The research was better because our experience informed almost all aspects of it. We helped make sure the research was relevant to the real world. |
| Total Score | 20 /30 |  |

##### Did the lived/living experience engagement process have any negative impacts on the research project?

☒ No

☐ Yes. Please describe:

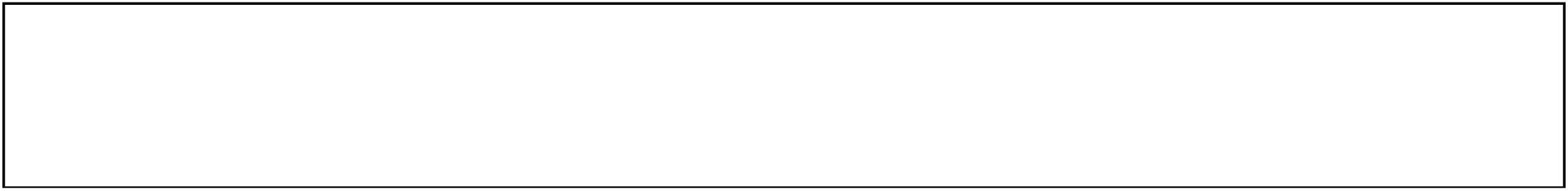

##### METRE Diagram

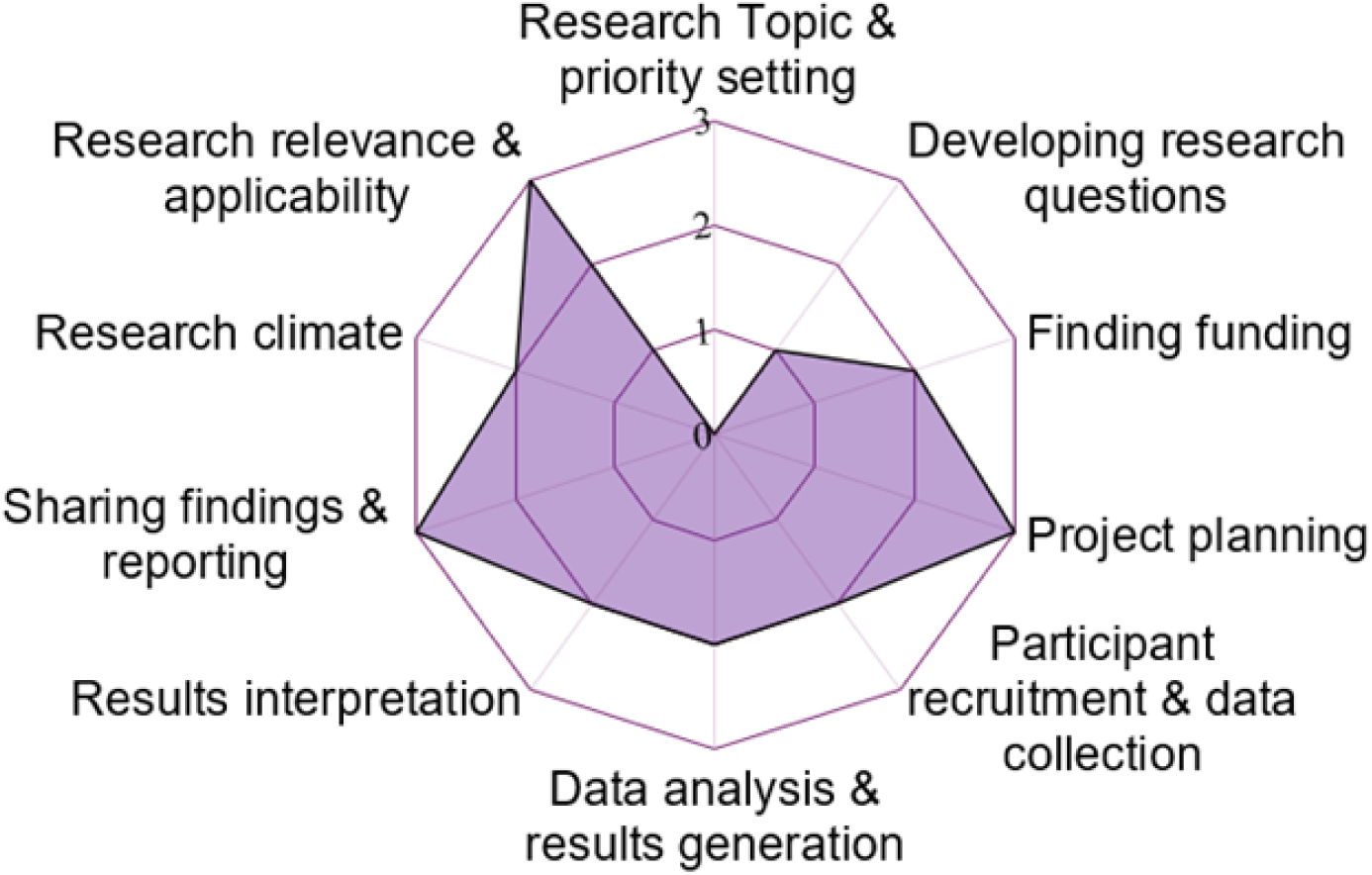

#### Area of Impact & Descriptions of Ratings

##### 1. Research topic & priority setting

This section refers to lived/living experience and caregiver engagement in formally or informally deciding what topics the research should be addressing (i.e., topic brainstorming, priority setting).

|  |  |
| --- | --- |
| **0** | There was **no impact** if people with lived/living experience and caregivers were not engaged in this stage or if their feedback was not taken into account. |
| **1** | There was a **small positive impact** if people with lived/living experience and caregiver feedback had limited impact on pre-established research topics or priorities. |
| **2** | There was a **medium positive impact** if people with lived/living experience and caregivers changed or refined pre-established research topics or priorities. |
| **3** | There was a **large positive impact** if people with lived/living experience and caregivers worked together with the academic researchers and staff to develop research topics or priorities early in the research process. |

| Your rating | Please describe |
| --- | --- |
| ☐ 0. No impact<br>☐ 1. Small positive impact<br>☐ 2. Medium positive impact<br>☐ 3. Large positive impact |  |

##### 2. Developing research questions

This section refers to lived/living experience and caregiver engagement in developing or refining the specific research questions that should be asked within a project.

|  |  |
| --- | --- |
| **0** | There was **no impact** if people with lived/living experience and caregivers were not engaged in this stage or if their feedback was not taken into account. |
| **1** | There was a **small positive impact** if people with lived/living experience and caregiver feedback led to minor adjustments to the research questions previously developed by academic researchers and staff. |
| **2** | There was a **medium positive impact** if people with lived/living experience and caregivers changed or improved the research questions previously developed by academic researchers and staff. |
| **3** | There was a **large positive impact** if people with lived/living experience and caregivers worked together with the academic researchers and staff to develop new research questions. |

| Your rating | Please describe |
| --- | --- |
| ☐ 0. No impact<br>☐ 1. Small positive impact<br>☐ 2. Medium positive impact<br>☐ 3. Large positive impact |  |

##### 3. Finding funding

This section refers to lived/living experience and caregiver engagement in the process of finding funding to support a project, regardless of whether the process was successful (e.g., institutional support, grant application, letter of support).

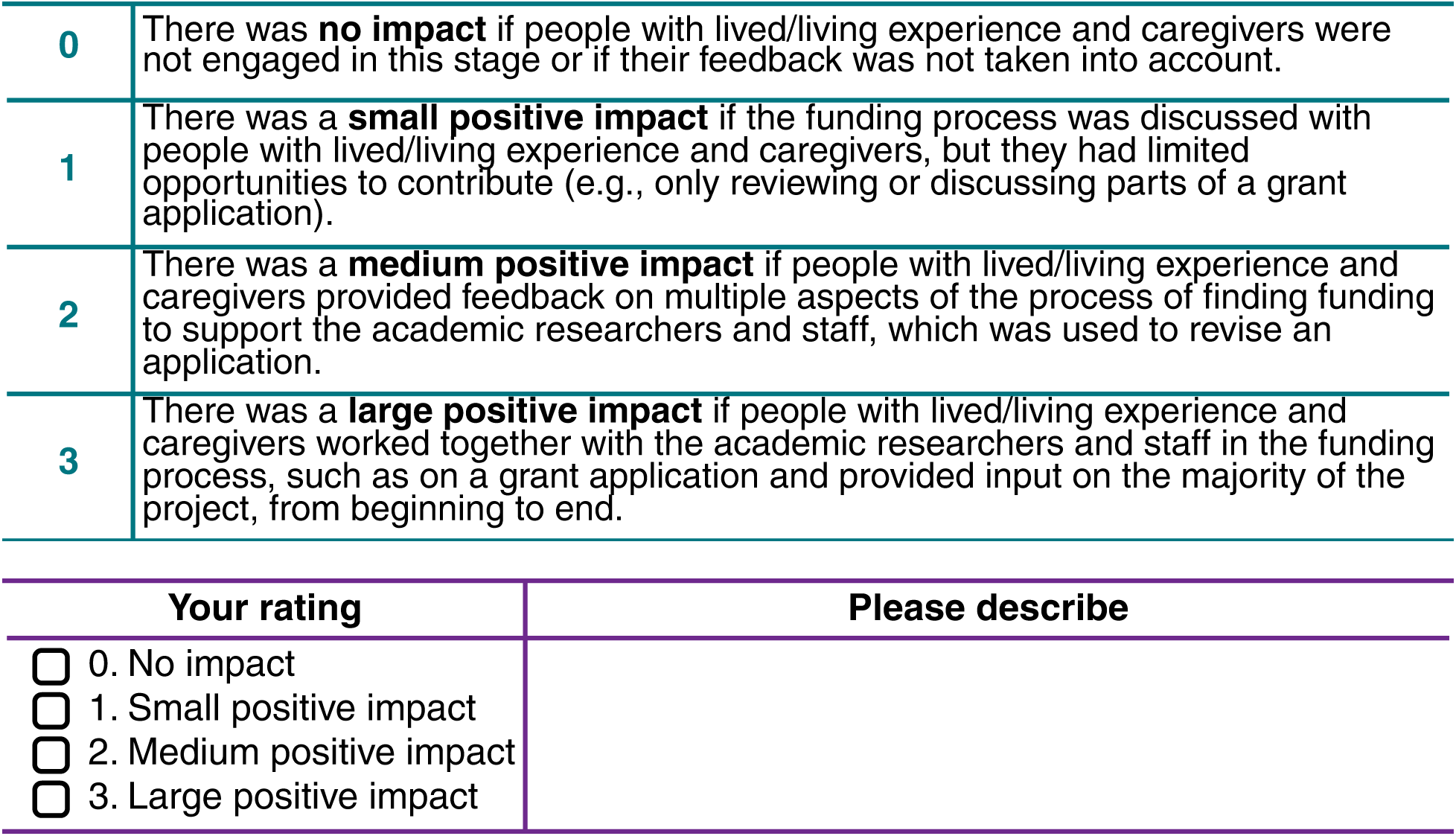

|  |  |
| --- | --- |
| 0 | There was **no impact** if people with lived/living experience and caregivers were not engaged in this stage or if their feedback was not taken into account. |
| 1 | There was a **small positive impact** if the funding process was discussed with people with lived/living experience and caregivers, but they had limited opportunities to contribute (e.g., only reviewing or discussing parts of a grant application). |
| 2 | There was a **medium positive impact** if people with lived/living experience and caregivers provided feedback on multiple aspects of the process of finding funding to support the academic researchers and staff, which was used to revise an application. |
| 3 | There was a **large positive impact** if people with lived/living experience and caregivers worked together with the academic researchers and staff in the funding process, such as on a grant application and provided input on the majority of the project, from beginning to end. |

| Your rating | Please describe |
| --- | --- |
| ☐ 0. No impact |  |
| ☐ 1. Small positive impact |  |
| ☐ 2. Medium positive impact |  |
| ☐ 3. Large positive impact |  |

##### 4. Project planning

This section refers to lived/living experience and caregiver engagement in the various planning steps leading up to the start of the project, such as preparing ethics board applications, drafting project materials, and developing project procedures

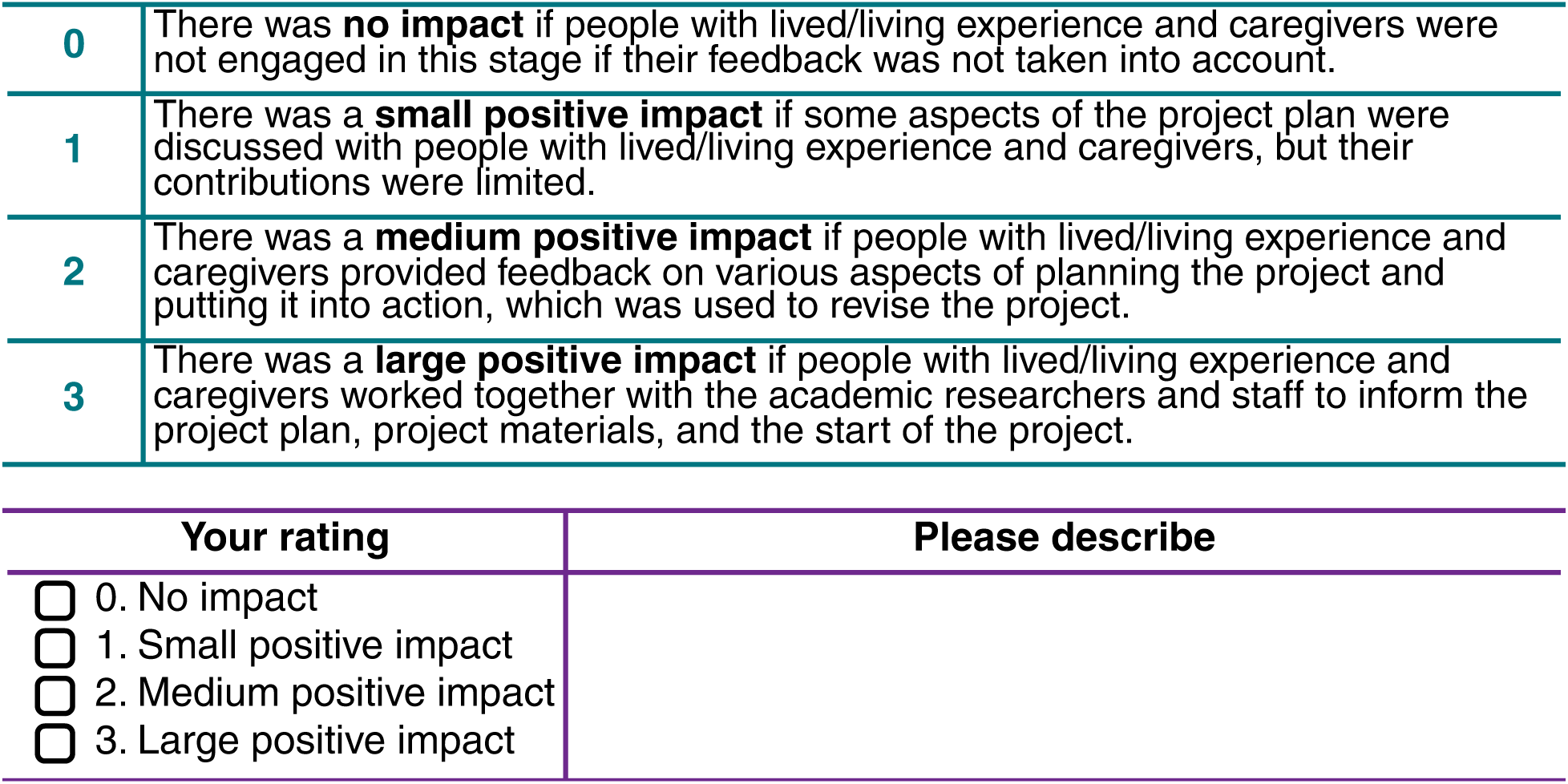

##### 5. Participant recruitment & data collection

This section refers to lived/living experience and caregiver engagement in the development of participant recruitment approaches or materials and the collection of data, whether the data were collected from people or from another source (e.g., literature review, animal models).

|  |  |
| --- | --- |
| 0 | There was **no impact** if people with lived/living experience and caregivers were not engaged in this stage or if their feedback was not taken into account. |
| 1 | There was a **small positive impact** if people with lived/living experience and caregivers made suggestions on the recruitment approaches, materials, or data collection process, which led to some improvements in the project. |
| 2 | There was a **medium positive impact** if people with lived/living experience and caregivers improved the recruitment and data collection process, such as by providing feedback on participant-facing materials, identifying diverse recruitment sources in their network, or suggesting data collection approaches. |
| 3 | There was a **large positive impact** if people with lived/living experience and caregivers were actively engaged in the recruitment planning process, creating visually appealing and accessible materials, directly recruiting participants, or personally collecting data. |

| Your rating | Please describe |
| --- | --- |
| ☐ 0. No impact<br>☐ 1. Small positive impact<br>☐ 2. Medium positive impact<br>☐ 3. Large positive impact |  |

##### 6. Data analysis & results generation

This section refers to lived/living experience and caregiver engagement in the process of analyzing the data or otherwise generating results from the project.

|  |  |
| --- | --- |
| 0 | There was **no impact** if people with lived/living experience and caregivers were not engaged in this stage or if their feedback was not taken into account. |
| 1 | There was a **small positive impact** if people with lived/living experience and caregivers had some limited engagement in analyzing the data or generating the results, such as reviewing data to inform the analysis. |
| 2 | There was a **medium positive impact** if people with lived/living experience and caregivers contributed to this stage by asking specific questions about the data to improve the analyses or by guiding decisions about the data, which was used to revise the analyses or generate results. |
| 3 | There was a **large positive impact** if people with lived/living experience and caregivers were actively engaged in collaborative data analysis sessions or personally generated the project's results. |

| Your rating | Please describe |
| --- | --- |
| ☐ 0. No impact<br>☐ 1. Small positive impact<br>☐ 2. Medium positive impact<br>☐ 3. Large positive impact |  |

##### 7. Results interpretation

This section refers to lived/living experience and caregiver engagement in understanding the findings, e.g., interpreting and providing context to the results.

|  |  |
| --- | --- |
| 0 | There was **no impact** if people with lived/living experience and caregivers were not engaged in this stage or if their feedback was not taken into account. |
| 1 | There was a **small positive impact** if people with lived/living experience and caregivers were engaged to some degree in interpreting the results, such as confirming whether or not the findings resonated with their experiences. |
| 2 | There was a **medium positive impact** if people with lived/living experience and caregivers contributed to this stage by suggesting, refining, or validating interpretations, identifying limitations, and providing additional context or clarifications. |
| 3 | There was a **large positive impact** if people with lived/living experience and caregivers provided new perspectives on the findings and their limitations and identified new, relevant implications for the results. |

| Your rating | Please describe |
| --- | --- |
| ☐ 0. No impact<br>☐ 1. Small positive impact<br>☐ 2. Medium positive impact<br>☐ 3. Large positive impact |  |

##### 8. Sharing findings & reporting

This section refers to lived/living experience and caregiver engagement in the process of sharing the findings of the project, such as reporting on the findings through manuscripts, presentations, tools for the community, or policy outputs.

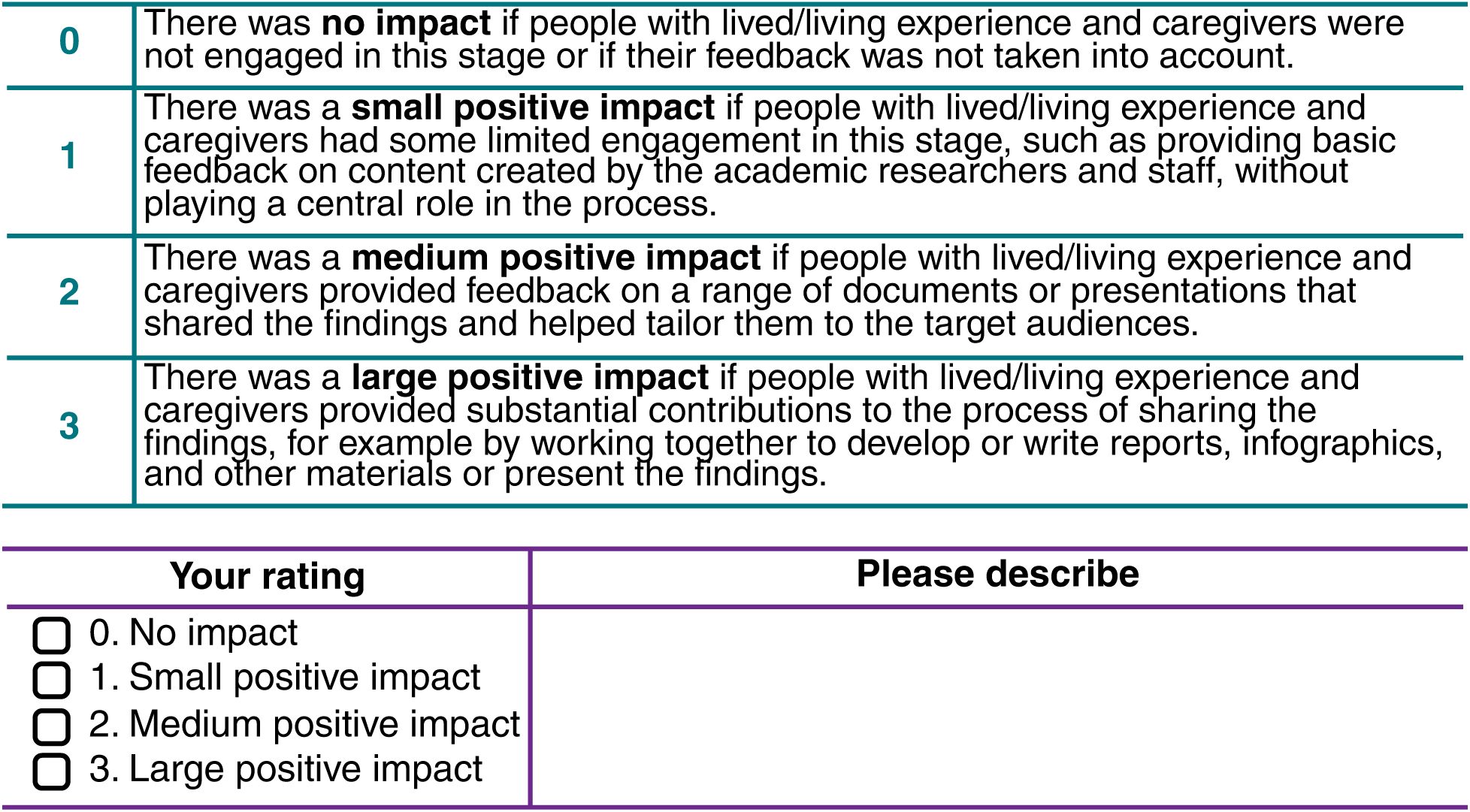

|  |  |
| --- | --- |
| 0 | There was **no impact** if people with lived/living experience and caregivers were not engaged in this stage or if their feedback was not taken into account. |
| 1 | There was a **small positive impact** if people with lived/living experience and caregivers had some limited engagement in this stage, such as providing basic feedback on content created by the academic researchers and staff, without playing a central role in the process. |
| 2 | There was a **medium positive impact** if people with lived/living experience and caregivers provided feedback on a range of documents or presentations that shared the findings and helped tailor them to the target audiences. |
| 3 | There was a **large positive impact** if people with lived/living experience and caregivers provided substantial contributions to the process of sharing the findings, for example by working together to develop or write reports, infographics, and other materials or present the findings. |

| Your rating | Please describe |
| --- | --- |
| ☐ 0. No impact<br>☐ 1. Small positive impact<br>☐ 2. Medium positive impact<br>☐ 3. Large positive impact |  |

##### 9. Research climate

This section refers to the way lived/living experience and caregiver engagement influenced the research climate, such as relationships, power sharing, and ways of working within the research team.

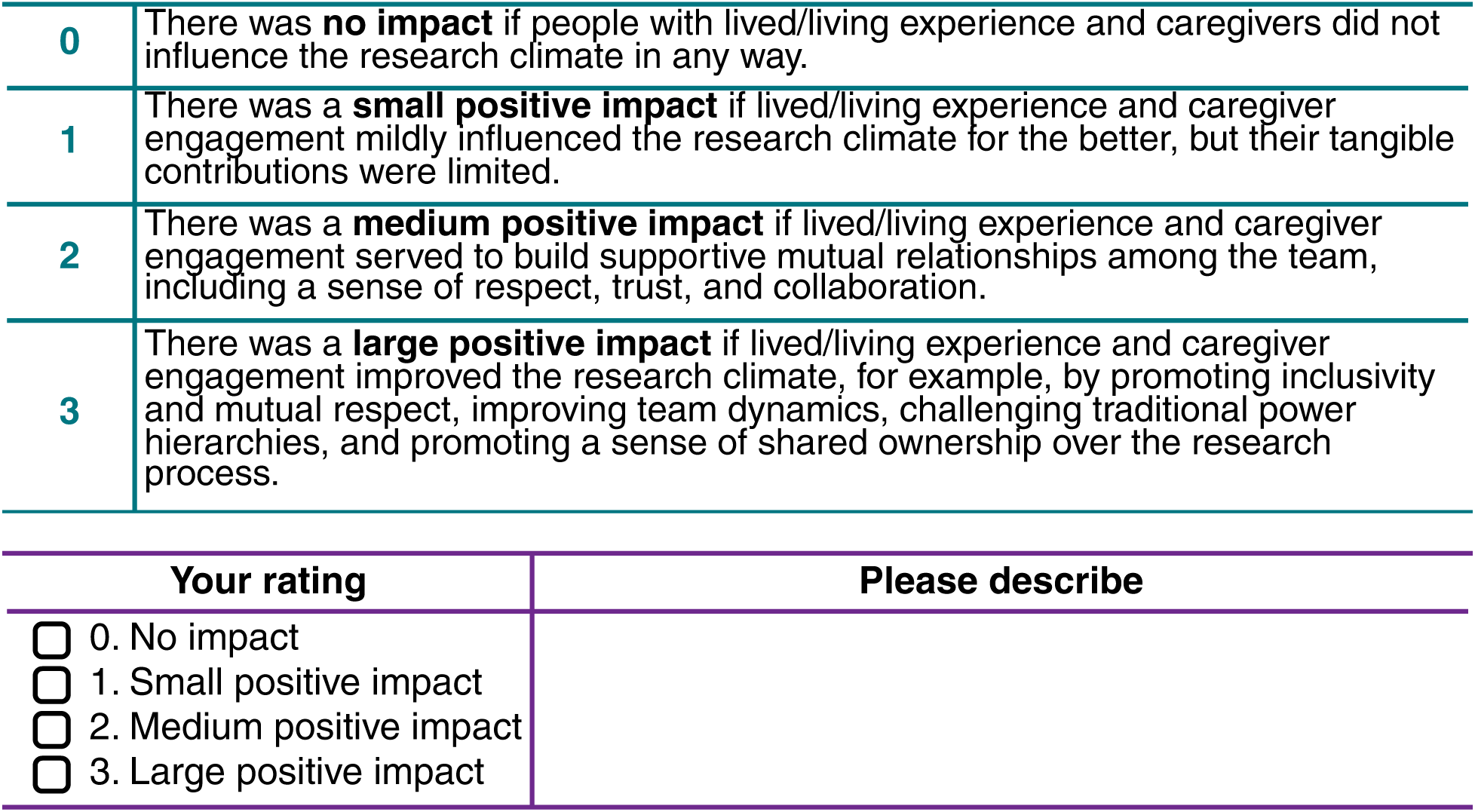

|  |  |
| --- | --- |
| 0 | There was **no impact** if people with lived/living experience and caregivers did not influence the research climate in any way. |
| 1 | There was a **small positive impact** if lived/living experience and caregiver engagement mildly influenced the research climate for the better, but their tangible contributions were limited. |
| 2 | There was a **medium positive impact** if lived/living experience and caregiver engagement served to build supportive mutual relationships among the team, including a sense of respect, trust, and collaboration. |
| 3 | There was a **large positive impact** if lived/living experience and caregiver engagement improved the research climate, for example, by promoting inclusivity and mutual respect, improving team dynamics, challenging traditional power hierarchies, and promoting a sense of shared ownership over the research process. |

| Your rating | Please describe |
| --- | --- |
| ☐ 0. No impact<br>☐ 1. Small positive impact<br>☐ 2. Medium positive impact<br>☐ 3. Large positive impact |  |

##### 10. Research relevance & applicability

This section refers to the overall impact of lived/living experience and caregiver engagement on how relevant the research is and how much it applies to real-life needs or experiences.

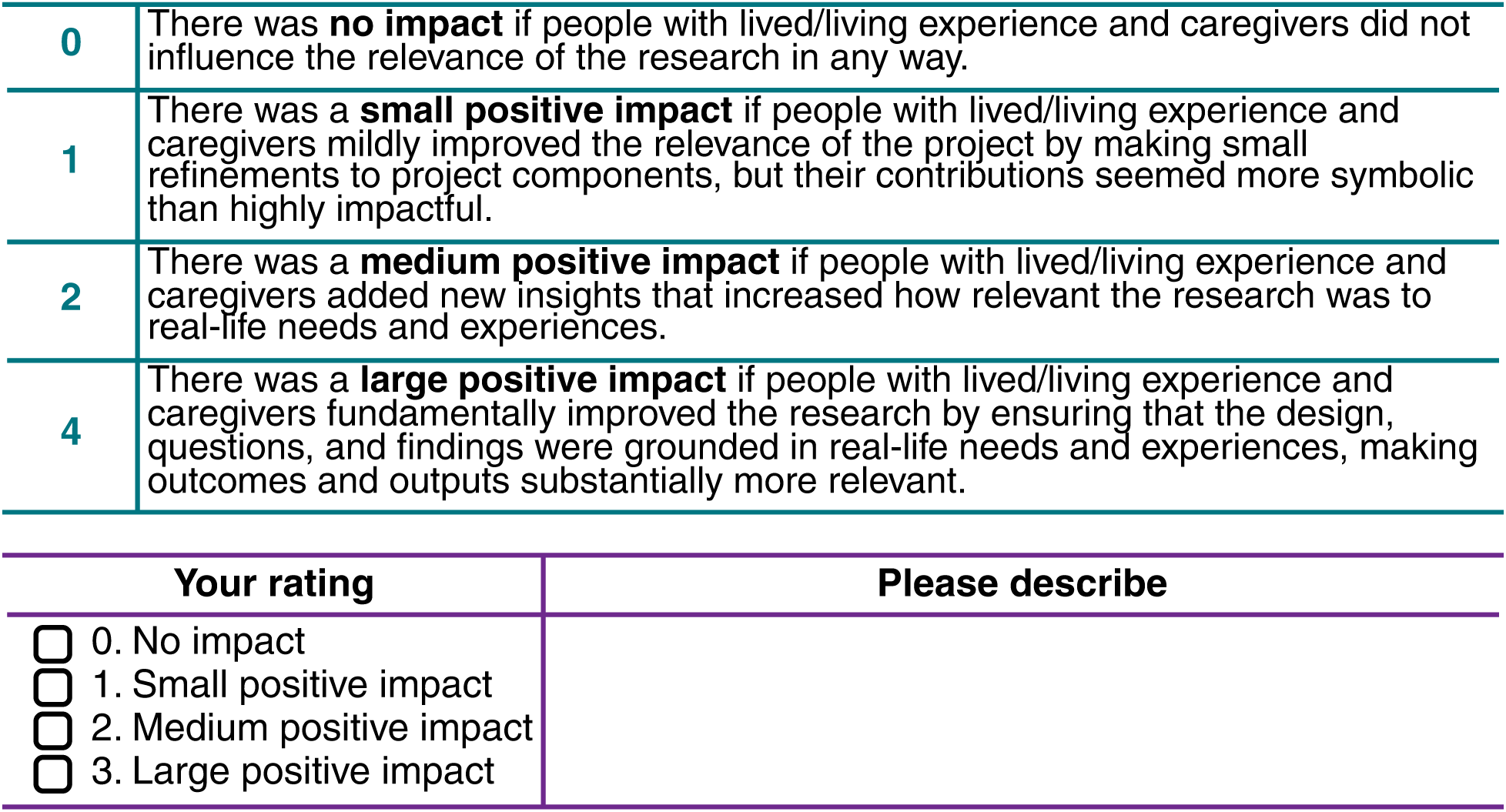

###### Did the lived/living experience engagement process have any negative impacts on the research project?

No

Yes. Please describe:

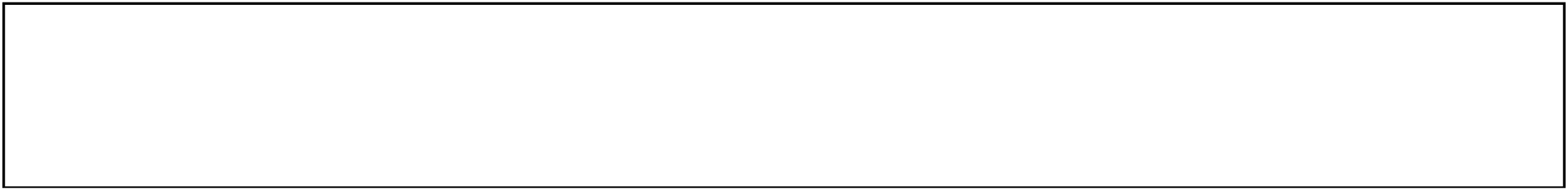

#### The Diagram

Once you’ve finished scoring the factors, open the **<u>Excel spreadsheet</u>** and enter the scores you’ve identified for each of the 10 areas of impact. Then, view the diagram that shows a picture of how much impact engagement has had across the domains of your project. The more coverage there is of the diagram, the more impact engagement has had on your project.

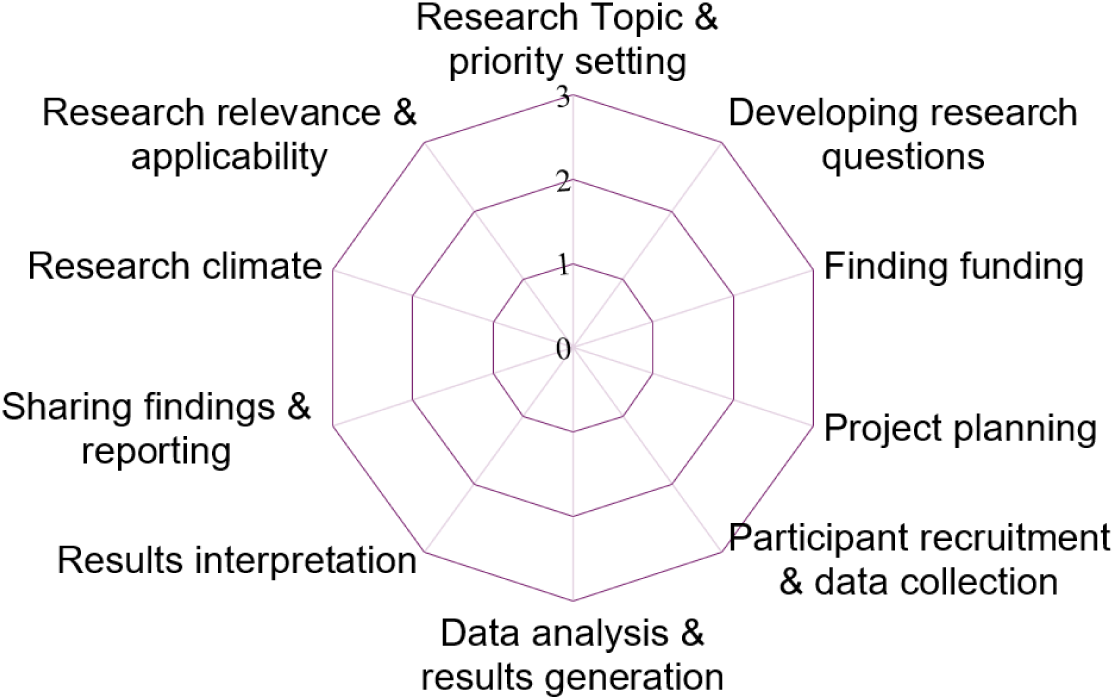

#### The METRE Scale

##### I am a

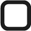 Lived experience advisor, collaborator, co-researcher
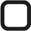 Family/caregiver advisor, collaborator, co-researcher
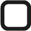 Scientist or academic investigator with a research institution
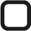 Research staff or academic trainee

###### Please use the scale below

0 No impact
1 Small positive impact
2 Medium positive impact
3 Large positive impact

| Area of Impact | Score | Describe |
| --- | --- | --- |
| 1. Research topic & priority setting |  |  |
| 2. Developing research questions |  |  |
| 3. Finding funding |  |  |
| 4. Project planning |  |  |
| 5. Participant recruitment & data collection |  |  |
| 6. Data analysis & results generation |  |  |
| 7. Results interpretation |  |  |
| 8. Sharing findings & reporting |  |  |
| 9. Research climate |  |  |
| 10. Research relevance & applicability |  |  |
| **Total Score** | **/30** |  |

##### Did the lived/living experience engagement process have any negative impacts on the research project?

No

Yes. Please describe:

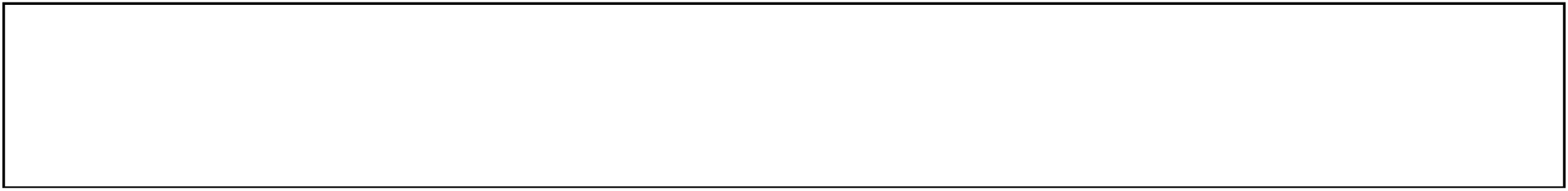

##### The Diagram

Once you’ve finished scoring the factors, open the **<u>Excel spreadsheet</u>** and enter the scores you’ve identified for each of the 10 areas of impact. Then, view the diagram that shows a picture of how much impact engagement has had across the domains of your project. The more coverage there is of the diagram, the more impact engagement has had on your project.

## Appendix D-2. METRE Scoring and Diagram

The Excel-based scoring tool for the METRE scale is provided as a supplementary file (Appendix D-2. METRE scoring - diagram Ver 2.0.xlsx). Respondent can enter item responses into the spreadsheet, which will automatically calculate the total score and generate visual summaries.

